# Prognostic Impact of Early Lactate Trajectory Among Patients Admitted with Cardiogenic Shock

**DOI:** 10.64898/2026.05.14.26353259

**Authors:** César Caraballo, Angela M Victoria-Castro, Aniket S. Rali, Eric J. Hall, Israel Safiriyu, Jason N. Katz, Ann Gage, Andrew P. Notarianni, David M. Dudzinski, Carlos Alviar, Guido Tavazzi, P. Elliott Miller

**Author notes:** **Corresponding Author**: P. Elliott Miller, MD, MHS, Section of Cardiovascular Medicine, Yale School of Medicine, New Haven, CT, 06517.

## Abstract

**Background:** The importance of lactate trajectory during the first day of cardiogenic shock is increasingly recognized. We aimed to assess the association between admission-day lactate trajectory and in-hospital mortality, and to identify same-day interventions predictive of lactate clearance.

**Methods:** We analyzed adult patients admitted with cardiogenic shock between October 2015 and June 2023, using the Vizient® Clinical Data Base. Early lactate clearance was defined as lactate <2.5 mmol/L by the end of the admission day. We used multivariable logistic regression to assess the association between lactate change and in-hospital mortality, and to identify interventions associated with lactate clearance.

**Results:** Among 40,434 patients with cardiogenic shock, 30.1% achieved same-day lactate normalization, which was associated with lower in-hospital mortality (aOR 0.51; 95% CI 0.48–0.54). Lactate change showed the greatest prognostic importance, with observed mortality exceeding 80% among those with lactate increase >5 mmol/L regardless of baseline values. After adjustment, lactate change showed a positive exponential relationship with mortality, with aORs ranging from 0.25 (95% CI 0.23–0.27) for a -10 mmol/L change to 3.99 (95% CI 3.58–4.40) for a +10 mmol/L change. The intervention most strongly associated with early lactate clearance was pulmonary artery catheter (PAC; aOR 1.28 [95% CI 1.19–1.37]).

**Conclusions:** Nearly 1 in 3 patients with cardiogenic shock achieved early lactate clearance, which was associated with lower mortality. The magnitude of lactate change had profound prognostic implications regardless of the initial value. Among day 1 interventions, PAC use had the strongest association with lactate clearance.

**CLINICAL PERSPECTIVE:** *What is new?:*

- In this real-world cohort of patients with cardiogenic shock, nearly 1 in 3 achieved normal lactate values on the first day of admission.
- Early lactate clearance was associated with nearly half the odds of in-hospital death, but the magnitude of lactate change on admission day had the strongest prognostic value regardless of the initial lactate value.
- The intervention on the first day of admission most associated with early lactate clearance was pulmonary artery catheterization placement.

*What are the clinical implications?:*

- Both the normalization of and magnitude of change in lactate on the first day of admission offer prognostic information that can help guide risk stratification in patients with cardiogenic shock.
- Our findings are further support for the importance of early trajectory in patients with cardiogenic shock and highlight the importance of lactate trends in that assessment.

## INTRODUCTION

Cardiogenic shock is a common critical cardiac condition that, despite progress, still has high mortality rates.^1^ Initial lactate levels are often used as a key prognostic indicator as they are known to correlate with mortality.^2–6^ Recent studies have highlighted the potential of early lactate clearance, defined as substantial reduction or normalization of lactate values within 24 hours of admission, as a prognostic marker in cardiogenic shock.^7–10^ However, the relationship between metabolic trajectory and survival remains poorly characterized across the spectrum of disease severity. Specifically, it is unclear how prognosis is affected by direction and magnitude of change in lactate concentration as a continuum and if severe admission hyperlactatemia is irreversible, or if it can be mitigated by rapid normalization within the first 24 hours. Furthermore, data regarding therapeutic interventions associated with successful lactate clearance are scarce.

Understanding the real-world association between the trajectory of change in lactate during the first day of admission and mortality may provide important prognostic information by allowing tailoring of interventions during initial management. If early lactate trajectory is a strong modifier of baseline risk, it may improve the prognostication based on initial lactate levels. In addition, if extreme admission lactate levels confer high mortality regardless of subsequent clearance, this would suggest a threshold of increased risk that reframes the therapeutic approach, for example helping identify earlier those who may benefit the most (or least) from mechanical circulatory support (MCS). Additionally, identifying therapeutic predictors of clearance could generate hypotheses to optimize cardiogenic shock management.

Therefore, in this multicenter observational cohort study, we leveraged real-world data from more than 40,000 patients with cardiogenic shock across the US to estimate the association between early lactate clearance and in-hospital mortality. We also aimed to evaluate the prognostic value of same-day lactate change as a continuous variable, across all initial lactate values, and to identify admission-day interventions associated with early metabolic recovery.

## METHODS

### Data source

We used de-identified patient-level data from Vizient® Clinical Data Base, an analytic platform for performance improvement that includes >1,000 hospitals across the United States, which encompasses nearly 97% of the nation’s Academic Medical Centers.^11^ As data was deidentified, it was deemed exempt from the Yale University Institutional Review Board review. Data from the Vizient Clinical Data Base used by permission of Vizient, Inc. All rights reserved.

### Study population

We included data from all adult patients (age ≥18 years) with a diagnosis of cardiogenic shock (ICD-10 code R57.0) admitted from October 1, 2015, to June 30, 2023, who had elevated serum lactate on admission (defined as the first lactate ≥2.5 mmol/L) and at least one same-day subsequent lactate value. We excluded patients with concomitant sepsis diagnosis, those who presented after an outside-hospital cardiac arrest, those with unknown discharge disposition or who were transferred to another hospital, and those with lactate values >99^th^ percentile (extreme outliers).

### Outcomes and variables of interest

Our primary outcome was in-hospital mortality. Exposure variables of interest were early lactate clearance, defined as an initially elevated serum lactate (≥2.5 mmol/L) with the last same-day measured lactate <2.5 mmol/L (achieving same-day normal levels), and change in lactate during the first day of admission (delta between the first and the last lactate measurement on day of admission). Day of admission was defined as the first calendar day of hospitalization.

Our secondary outcome was early lactate clearance, defined as above, and the exposure variables were interventions on day one of admission: pulmonary artery catheter (PAC), left heart catheterization, percutaneous coronary intervention (PCI), intra-aortic balloon pump (IABP), Impella device, extracorporeal membrane oxygenation (ECMO), non-invasive ventilation, invasive mechanical ventilation, renal replacement therapy, and use of inotropes or vasopressors including norepinephrine, epinephrine, vasopressin, dopamine, milrinone, dobutamine, and phenylephrine. **Table S1** lists the International Classification of Diseases, Tenth Revision, Clinical Modification (ICD-10-CM) codes used to identify procedures.

We also included demographic data such as age (years), race and ethnicity (categorized as non-Hispanic Black, non-Hispanic White, non-Hispanic other, and Hispanic), sex (male, female), primary payer (commercial/private, Medicaid, Medicare, or other), and income quartile (<$47,001, $47,001–60,000, $60,001–80,000, or >$80,001); calculated Society for Cardiovascular Angiography & Intervention (SCAI) shock stages (A, B, C, D, E);^12^ comorbidities present at admission including Charlson Comorbidity Index, diabetes mellitus, peripheral vascular disease, chronic kidney disease, end-stage renal disease, rheumatic disease, chronic pulmonary disease, HIV/AIDS, cancer, chronic liver disease, coronary artery disease, dementia, dyslipidemia, smoking, prior myocardial infarction, prior PCI, prior coronary artery bypass graft (CABG), heart failure, history of left ventricular assist device (LVAD), history of heart transplant; laboratory results from day 1 of admission: WBC, hemoglobin, platelets, sodium, glucose, creatinine, blood urea nitrogen (BUN), estimated glomerular filtration rate (eGFR), N-terminal prohormone of brain natriuretic peptide (NT-proBNP), total bilirubin, aspartate aminotransferase (AST), alanine aminotransferase (ALT), albumin, serum pH, and bicarbonate); and hospital- and admission-level data used included US region (South, Midwest, West, Northeast), bed size (<125 beds, 125–300 beds, 301–500 beds, or >500 beds), weekday of admission, and teaching hospital or not. Lactate data, and other laboratory values, include sequence data (first, second, third, etc.) and are date-stamped to the calendar day of admission (day 1, day 2, etc.).

### Statistical analysis

We described our data stratified by early lactate clearance (cleared on day of admission vs. not cleared on day of admission). We described the study population’s sociodemographic, hospital and admission characteristics, baseline comorbidities, initial lab values, and day-one interventions. Continuous variables were described with mean and standard deviation if normally distributed or median and interquartile range (IQR) if not. Categorical variables were described with frequency and percentages. Categorical variables were compared using the Chi-square test. Numerical variables were compared using t-test or Wilcoxon rank-sum test, depending on their distribution.

We first described crude mortality rates by early lactate clearance status followed by bivariate logistic regression to report odds ratios for in-hospital mortality. We then used multivariable logistic regression using in-hospital mortality as the outcome and early lactate clearance as the explanatory variable, including demographic, comorbidities, laboratory data (hemoglobin, total bilirubin, AST, ALT, creatinine, first lactate), and hospital-level data as covariables to adjust for confounders. To visualize the combined association between initial lactate concentration, its same-day change, and in-hospital mortality, we plotted a heatmap using hexagonal binning with a color gradient representing the observed in-hospital mortality rate. In addition, to assess the risk association in a more continuous manner, we used marginal standardization using a logistic regression model with the same covariates (fixed at their sample means) as the model described above but replacing the main exposure variable of interest with the absolute same-day change in lactate (mmol/L). Using this approach, we estimated the predicted odds of mortality across a spectrum of lactate changes (–10 to +10 mmol/L). These predicted values were normalized to a reference group of no change (same-day change in lactate = 0 mmol/L) to obtain adjusted odds ratios (aOR) and 95% confidence intervals. For consistency with other studies on this topic,^9,13–15^ we also used Kaplan-Meier curves and multivariable Cox proportional hazards regression models to evaluate the association between early lactate clearance and mortality within 30 days of admission.

For the secondary outcome, we used multivariable logistic regression using early lactate clearance as the outcome and day one interventions as the main explanatory variables of interest. Demographic, hospital and admission characteristics, laboratory data (first lactate, hemoglobin, total bilirubin, AST, ALT, creatinine) and comorbidities were included in the model as covariates.

We performed sensitivity analyses. For the main outcome, since we only have calendar day-level data, we expanded the lactate clearance period to the second calendar day of admission. We also performed a sensitivity analysis using a higher lactate threshold, using 4 mmol/L instead of 2.5 mmol/L.

Statistical significance was set at P<0.05 for all analyses. Odds ratios were reported with 95% confidence intervals. All analyses were performed in Stata 17 SE (Statacorp, College Station, TX).

## RESULTS

### Population admission characteristics

We included data from 40,434 individuals with cardiogenic shock who met the inclusion criteria (**Figure S1**). Of these, 12,157 (30.1%) achieved normal lactate levels on the last measurement on the admission day (early lactate clearance). When compared with those who did not clear lactate on day 1, those who achieved early lactate clearance had comparable sociodemographic characteristics, although were slightly more likely to be Black and to be treated at a teaching or larger bed size hospital (P<0.001 for each; **Table 1**).

**Table 1.**
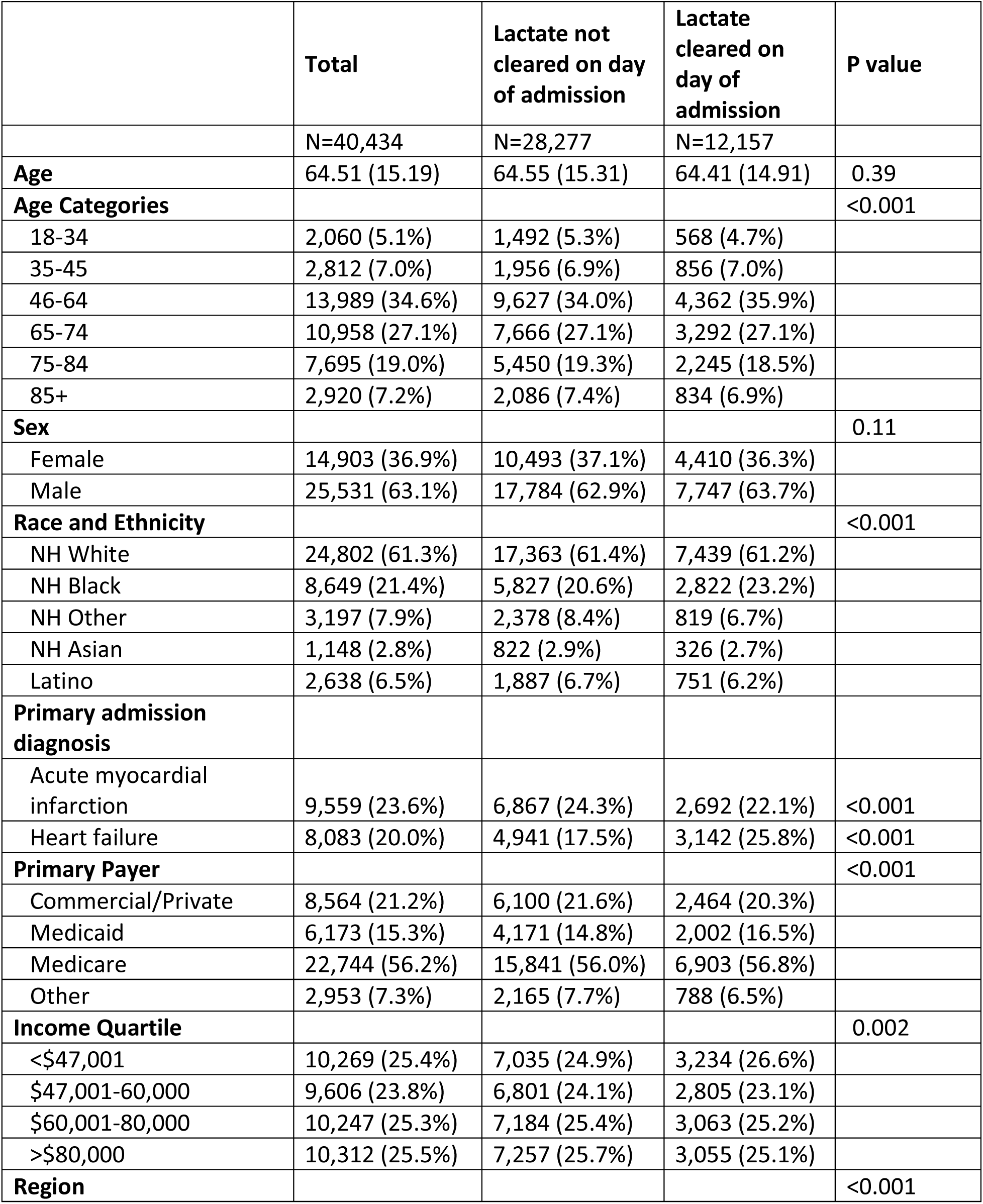
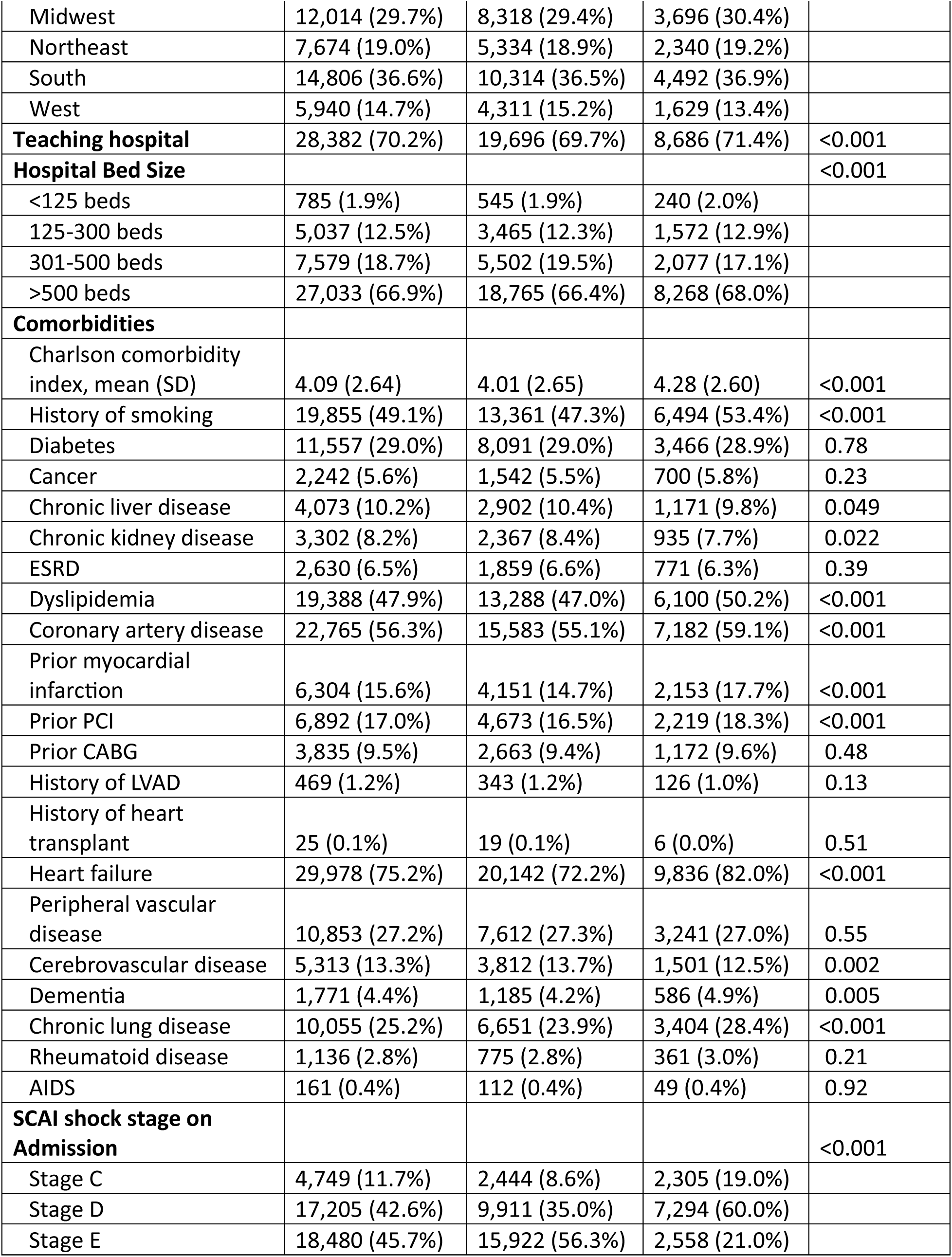
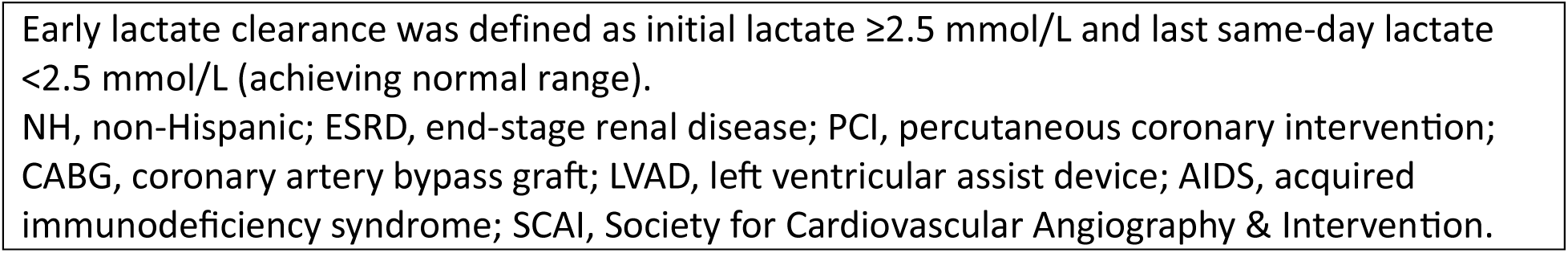
Baseline population sociodemographic characteristics and comorbidities among patients with cardiogenic shock and initial lactate elevation, stratified by early lactate clearance

When compared with those without early lactate clearance, those who achieved early lactate clearance had higher burden of chronic conditions, including heart failure and coronary artery disease, and higher Charlson comorbidity index score (P<0.001 for each; Table 1). However, they presented with lower acuity, characterized by lower SCAI shock stages and lower biomarkers of hypoperfusion. The median (IQR) first lactate was 6.0 (3.9–9.3) mmol/L among those who did not clear lactate on day 1, compared with 3.7 (2.9–5.4) mmol/L among those who did (P <0.001; **Table 2**). The distribution of admission lactate, last lactate, and lactate change is shown in **Figure S2**. Those who achieved early lactate clearance had lower creatinine levels (median [IQR] 1.50 [1.20–2.27] vs. 1.56 [1.12–2.36] mg/dL; P <0.001), lower NT-ProBNP (6630 [2195–17438] vs. 7644 [1885–21790] pg/mL; P=0.01), along with lower total bilirubin, AST, and ALT (P<0.001 for each; **Table 2**).

**Table 2.** Initial laboratory results among patients with cardiogenic shock and initial lactate elevation, stratified by early lactate clearance

|  | Total | Lactate not cleared<br>on day of admission | A Lactate cleared on<br>day of admission | P<br>value |
| --- | --- | --- | --- | --- |
| **Laboratory results** | N=40,434 | N=28,277 | N=12,157 |  |
| First lactate,<br>mmol/L | 5.10 (3.40-8.20) | 6.0 (3.90-9.30) | 3.70 (2.90-5.40) | <0.001 |
| Max lactate,<br>mmol/L | 6.40 (4.10-10.20) | 7.90 (5.10-11.70) | 4.0 (3.10-6.0) | <0.001 |
| Last lactate,<br>mmol/L | 3.61 (2.20-6.90) | 5.20 (3.50-8.70) | 1.80 (1.40-2.10) | <0.001 |
| WBC, 1000/μL | 13.97 (10.24) | 14.38 (10.38) | 13.01 (9.82) | <0.001 |
| Hemoglobin, g/dL | 12.20 (2.75) | 12.17 (2.78) | 12.27 (2.67) | <0.001 |
| Platelets, 1000/μL | 215.47 (104.63) | 210.57 (106.17) | 226.88 (100.04) | <0.001 |
| Sodium, mmol/L | 136.98 (6.06) | 137.28 (6.15) | 136.28 (5.77) | <0.001 |
| Glucose, mg/dL | 172.0 (122.0-255.0) | 176.0 (123.0-263.0) | 165.0 (122.0-236.0) | <0.001 |
| Creatinine, mg/dL | 1.53 (1.11-2.32) | 1.56 (1.12-2.36) | 1.50 (1.10-2.27) | <0.001 |
| BUN, mg/dL | 25.50 (17.0-42.0) | 25.0 (17.0-42.0) | 27.0 (17.0-44.0) | <0.001 |
| eGFR,<br>ml/min/1.73m<sup>2</sup> | 45.0 (27.0-60.0) | 44.0 (26.0-60.0) | 46.0 (28.0-60.0) | <0.001 |
| NT-proBNP,<br>pg/mL | 7283 (1994-20262) | 7644.0 (1885.0-21790.0) | 6630.0 (2195.0-17438.0) | 0.014 |
| Total bilirubin,<br>mg/dL | 0.90 (0.50-1.70) | 0.90 (0.50-1.80) | 0.80 (0.50-1.50) | <0.001 |
| AST, U/L | 533.61 (1563.86) | 625.77 (1749.05) | 322.68 (988.78) | <0.001 |
| ALT, U/L | 326.81 (860.12) | 375.03 (947.66) | 216.27 (599.55) | <0.001 |
| Albumin, g/L | 3.26 (0.67) | 3.21 (0.69) | 3.38 (0.61) | <0.001 |
| pH | 7.29 (7.18-7.38) | 7.28 (7.16-7.37) | 7.33 (7.24-7.40) | <0.001 |
| CO<sub>2</sub> (bicarb),<br>mmol/L | 19.72 (5.66) | 19.11 (5.75) | 21.12 (5.18) | <0.001 |
| Data are presented as mean (standard deviation) or median (percentile 25<sup>th</sup> – percentile 75<sup>th</sup>).<br>Early lactate clearance was defined as initial lactate ≥2.5 mmol/L and last same-day lactate <2.5 mmol/L (achieving normal range).<br>WBC, white blood cells; BUN, Blood urea nitrogen; eGFR, estimated glomerular filtration rate; NT-proBNP, N-terminal prohormone of brain natriuretic peptide; AST, aspartate aminotransferase;<br>ALT, alanine aminotransferase |  |  |  |  |

**Table 3.**
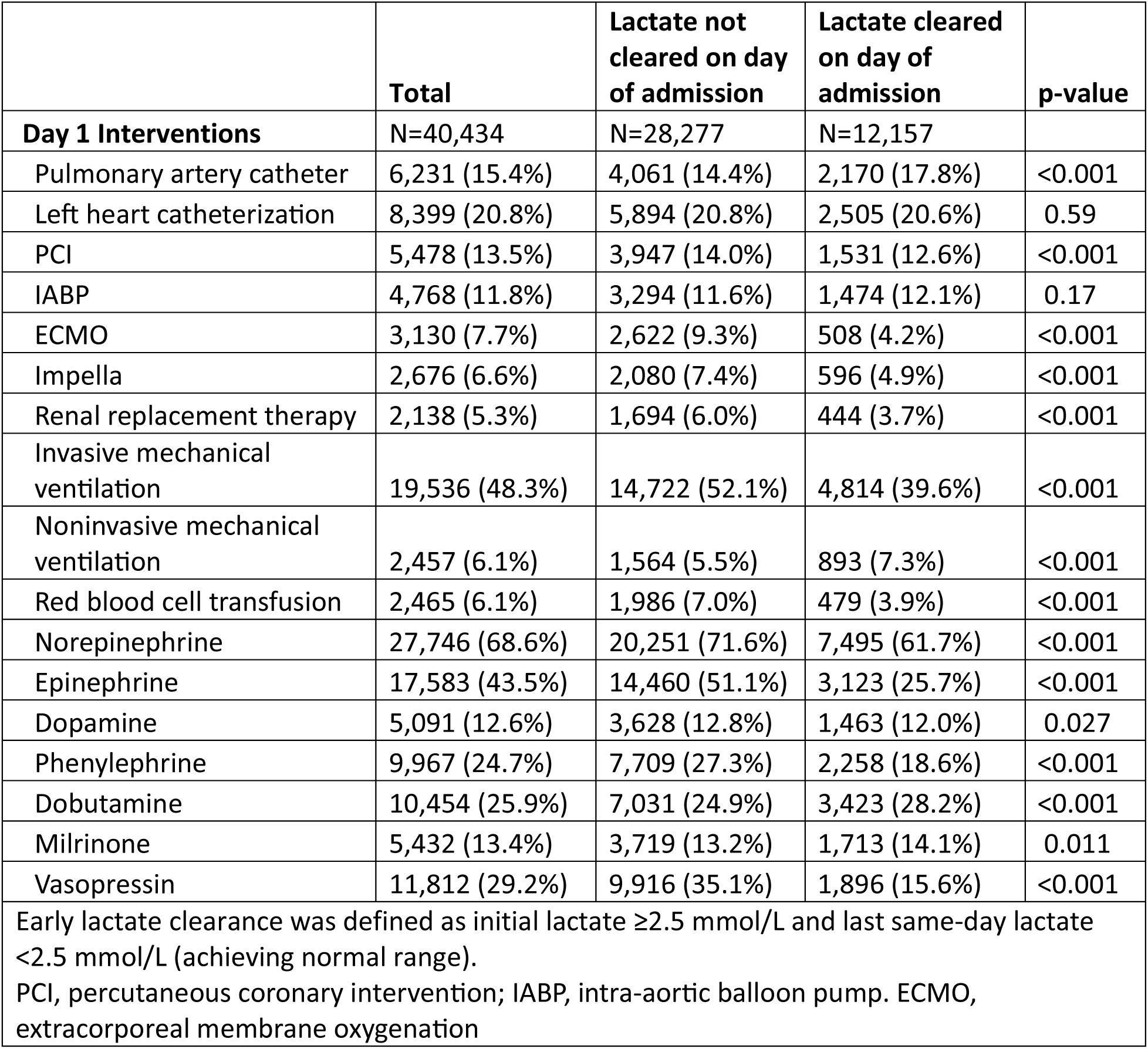
Admission day interventions among patient with cardiogenic shock and initial lactate elevation.

### Interventions on day of admission

In unadjusted analysis, those who achieved early lactate clearance had higher rates of PAC (17.8% vs. 14.4%; P<0.001), non-invasive mechanical ventilation (7.3% vs. 5.5%; P<0.001) use of milrinone (14.1% vs. 13.2%; P=0.01), and use of dobutamine (28.2% vs. 24.9%; P<0.001). In contrast, those who did not clear lactate were more likely to receive ECMO (9.3% vs. 4.2%), Impella (7.4% vs. 4.9%), renal replacement therapy (6.0% vs. 3.7%), mechanical ventilation (52.1% vs. 39.6%), and vasopressor support (P<0.001 for each;3).

### Early lactate clearance and in-hospital mortality

In-hospital mortality among those who achieved early lactate clearance was 22.8% compared to 47.9% (OR 0.32; 95% CI 0.31, 0.34) among those who did not. When adjusted for confounders, including the initial lactate level, early lactate clearance remained associated with lower in-hospital mortality (aOR 0.51; 95% CI 0.48, 0.54).

Lactate trajectory was associated with a substantial prognostic influence regardless of baseline values (**Figure 1 and Figure S3**). As an example, among patients with a severely elevated initial lactate (i.e., >10 mmol/L), a same-day reduction of >5 mmol/L resulted in observed mortality rates (31–50%) comparable to those who had mild initial elevations (2.5–5 mmol/L) and a same-day increase up to +5 mmol/L. In contrast, we found a gradient of increased mortality with any degree of lactate increase regardless of the initial value. Specifically, an increase of >5 mmol/L represented a shift towards observed mortality of 81-100% across nearly all initial lactate values.

**Figure 1.**
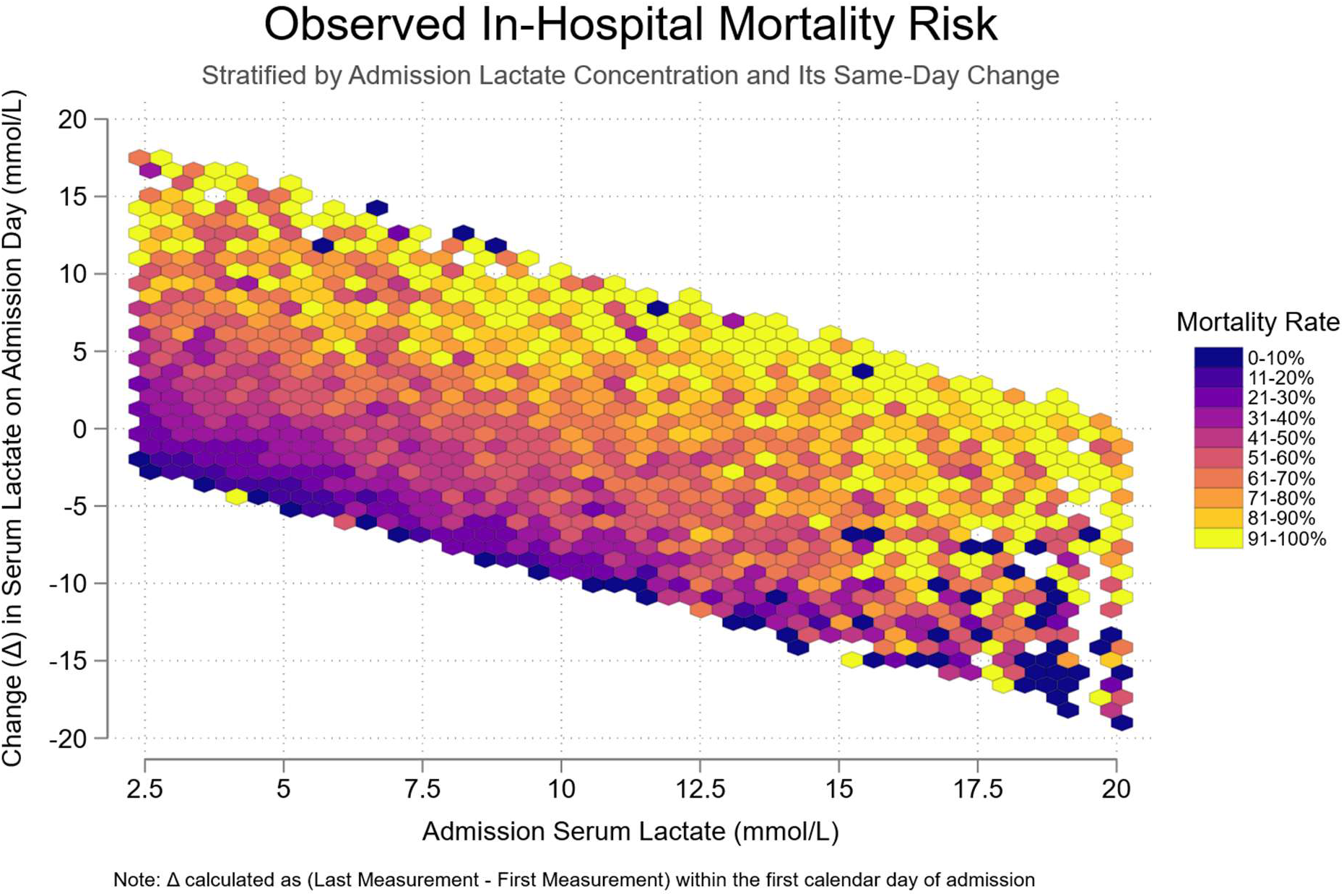
Heatmap of Observed In-Hospital Mortality Risk Among Patients with Cardiogenic Shock. In this heatmap, color intensity represents the observed mortality rate within each hexagonal bin across the bivariate distribution of admission lactate on the x-axis and same-day lactate change on the y-axis (each in mmol/L), ranging from 0-10% in dark purple to >90% in bright yellow.

After adjusting for confounders, an exponential relationship between lactate change and mortality persisted (**Figure 2 and Figure S4**). When compared with no change in lactate on the first day of admission, a net decrease in lactate was consistently protective, with reductions of >5 mmol/l more than halving the odds of mortality and reaching an aOR of 0.25 (95% CI: 0.23, 0.27) for a -10 mmol/L change. Similarly, a net increase was associated with an increased mortality, reaching an aOR of 3.99 (95% CI: 3.58, 4.40) for a +10 mmol/L change.

**Figure 2.**
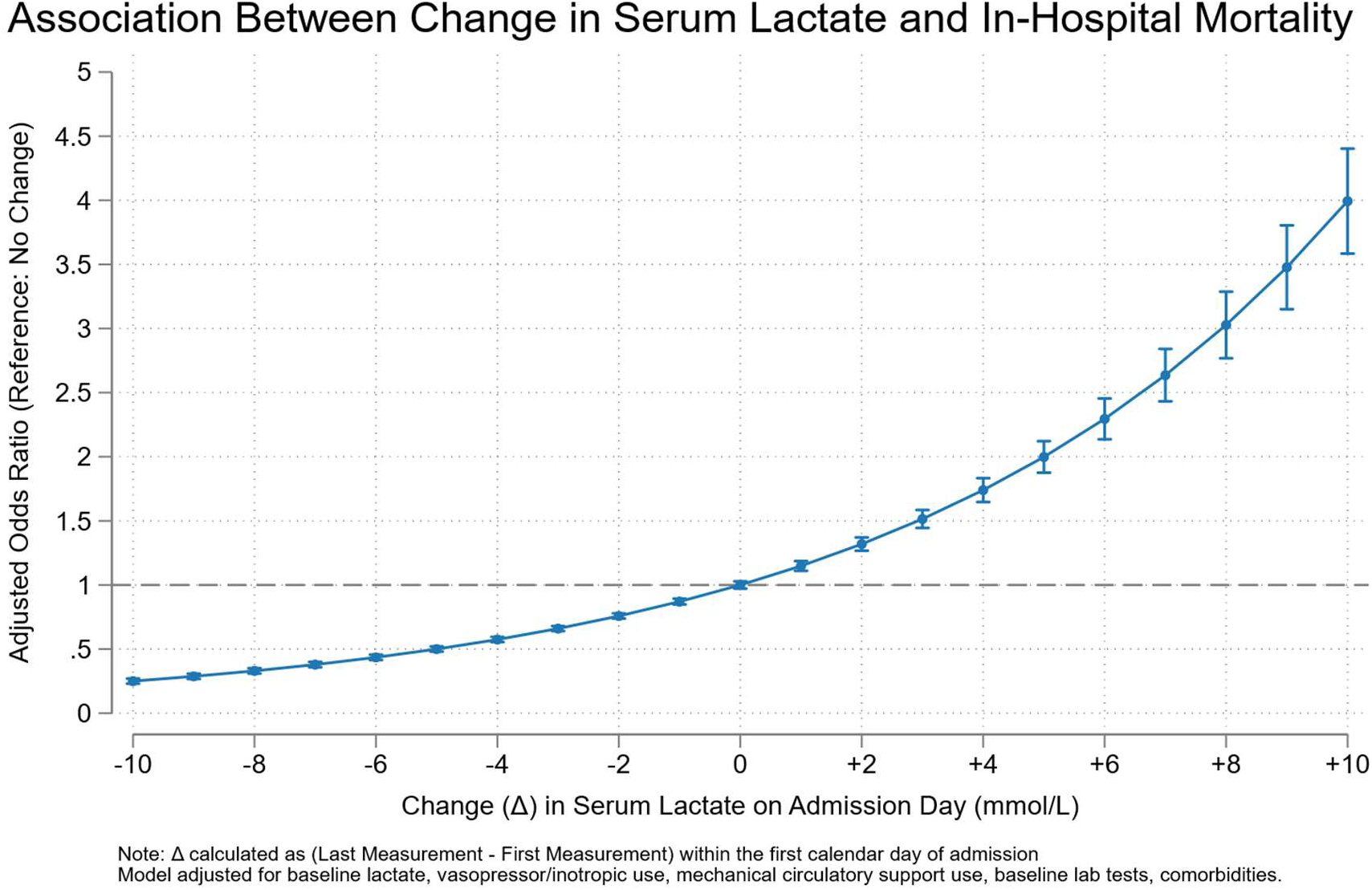
Association between change in serum lactate concentration and in-hospital mortality among patients with cardiogenic shock and initial serum lactate elevation. Odds ratios point estimates are represented with circles and error bars represent 95% confidence interval boundaries. Multivariable logistic regression included demographics, comorbidities, hospital/admission characteristics, labs (including first lactate level), and day 1 interventions (see methods section for details).

The association between early lactate clearance and in-hospital mortality persisted in a sensitivity analysis expanding the defined lactate clearance period up to the second calendar date of admission, which included 48,045 patients with initial lactate elevation and at least one subsequent lactate value in the period of day 1 and 2. The mortality rates were 60.8% among those who did not clear lactate by day 2 and 23.2% among those who did, with an adjusted mortality OR of 0.26 (95% CI 0.25, 0.27), and a steeper association between lactate change and mortality (**Figures S5 and S6**).

The main findings were also consistent in our sensitivity analysis increasing the threshold of lactate elevation from 2.5 to 4 mmol/L in which lactate clearance had an aOR 0.46 (95% CI 0.43, 0.50) for mortality (**Figures S7 and S8**).

In a survival time analysis, early lactate clearance was also associated with lower 30-day mortality, with an unadjusted HR of 0.43 (95% CI 0.41, 0.44) and an adjusted HR of 0.60 (95% CI 0.57, 0.63; **Figure S9**.).

### Interventions associated with early lactate clearance

After multivariable adjustment, the only intervention associated with successful same-day lactate clearance was PAC use (OR 1.28; 95% CI 1.19, 1.37). The remainder of interventions, including inotropes, vasopressors, and mechanical ventilation were either not significant or were associated with lower odds of clearance, likely reflecting higher disease severity in the non-clearance group (**Figure 3 and Table S2**).

**Figure 3.**
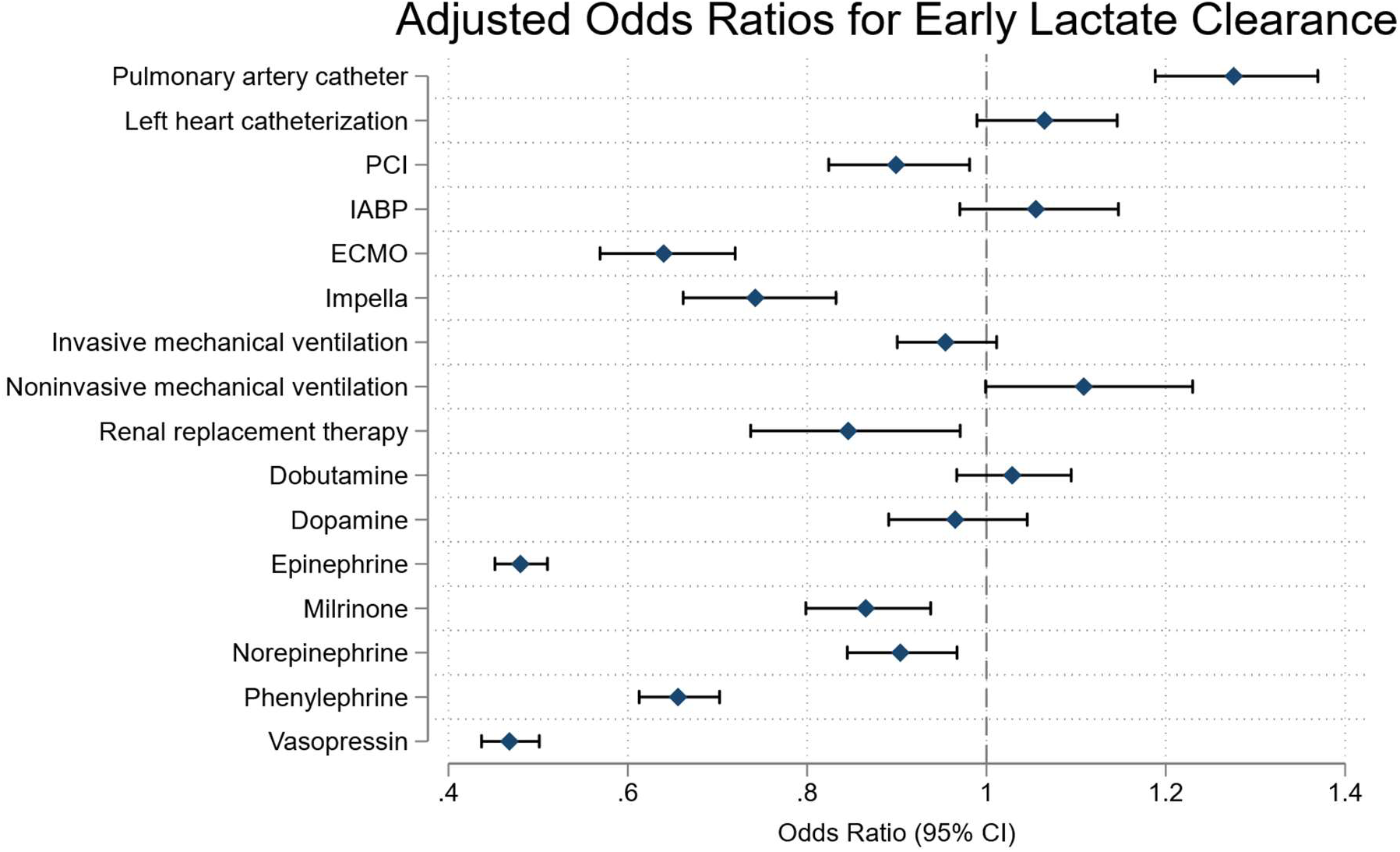
Association between interventions on day of admission and same-day lactate clearance among patients with cardiogenic shock. Odds ratio point estimates represented with circle, bars representing 95% CI. Multivariable logistic regression included demographics, comorbidities and hospital/admission characteristics (see methods section for details). Early lactate clearance was defined as initial lactate ≥2.5 mmol/L and last same-day lactate <2.5 mmol/L (achieving normal range). Abbreviations: PCI, percutaneous coronary intervention; IABP, intra-aortic balloon pump. ECMO, extracorporeal membrane oxygenation.

## DISCUSSION

In this multi-center observational study of over 40,000 patients hospitalized with cardiogenic shock, we found that early lactate clearance was a significant predictor of in-hospital survival. Moreover, the magnitude of change in lactate on the admission day was correlated with a substantial prognostic influence regardless of the initial lactate level, with observed mortality rates exceeding 80% among those with same-day serum lactate increase greater than 5–10 mmol/L, whereas those with decrease >5 mmol/L saw their risk significantly attenuated. In multivariable regression models, change in serum lactate concentration during day of admission had a robust exponential relationship with mortality. Notably, use of PAC was associated with greater odds of achieving early lactate clearance. These findings were consistent across several sensitivity analyses.

This study expands the literature in several ways. First, to our knowledge, this is the largest study that has shown the prognostic importance of early lactate clearance among patients with cardiogenic shock. Sample sizes from studies on this topic often range in the hundreds, with a meta-analysis including over 1200 patients,^13^ limiting such a granular assessment as ours. Second, we found that lactate trajectory modifies baseline risk overall and by initial lactate value, providing a fuller picture of the prognostic implication of same-day lactate trajectory. Those who had markedly elevated initial lactate had their mortality odds decreased to levels compared to that of those with mildly elevated levels by having a negative delta on same day. Conversely, patients with mild initial elevations who had marked increase in lactate concentration had risks comparable to those with higher initial lactate and minimal same-day change. Third, we used real-world patient-level data in a nationally representative database, with findings consistent with those of longitudinal registries, single-center observational studies, and secondary analyses of clinical trials.^7^ Fourth, in our secondary objective, we report that use of PAC is associated with early lactate clearance.

Our findings are consistent with existing literature. Smaller studies have consistently shown that lactate clearance during the first day of admission has a positive survival association.^1,15^ A smaller cohort of 628 patients with cardiogenic shock showed that early lactate clearance was associated with lower odds of in-hospital mortality (aOR 0.42, 95% CI 0.30–0.60),^8^ similar to our estimate. Regarding change over time, a smaller study using data from 171 patients with cardiogenic shock,^1^ found that those who did not survive had, on average, increasing or plateauing lactate during the first day of admission. In another study, Wu et al. found that, beyond specific values, increased time under elevated lactate values on day of admission had a similar association with in-hospital mortality.^14^ Similarly, Sundermeyer et al. described differences in survival based on short-interval lactate kinetics,^16^ although those with high initial lactate persisted with elevated risk of death regardless of 90-minute change. We found that, among our population, same-day change was indeed protective, with such a difference likely arising from the length of the interval between measurements.

Our study has important implications for clinical practice. Our findings further reinforce that serial lactate values on day of admission may be an important indicator of successful resuscitation and restoration of perfusion, potentially guiding decision-making. Even among those with the highest initial risk, capturing the trajectory of lactate concentration on the first day could inform with greater accuracy their evolving risk of death, rather than a single static value. We believe our findings are important both for clinicians prognosticating patients with cardiogenic shock and also for counseling families.

It is important to note the complexity of factors contributing to lactate production and clearance. Not all serum lactate elevation is secondary to global systemic hypoperfusion or tissue ischemia.^17,18^ In our study, for example, epinephrine was associated with lower odds of lactate clearance, but such an association could be mostly explained by its beta-adrenergic effect on metabolic pathways. Furthermore, in our study the population that did not achieve lactate clearance had worse indicators of liver and kidney function, which are the main mechanisms of serum lactate elimination. Physicians should remain attentive to situations such as these in which persistently elevated serum lactate may not represent failure of resuscitation.

Moreover, regarding hemodynamic monitoring, our findings suggest that use of PAC as part of standard management of cardiogenic shock may be beneficial. Much has been debated about the routine use of PAC among critically ill patients, with some results suggesting no benefit on mortality and increased complexity of care.^19–21^ However, the representation of patients with cardiogenic shock in these landmark studies was limited, and increasingly expert opinions have called for the use of PAC in patients with this condition.^22,23^ Although the use of PAC in patients with cardiogenic shock has decreased in frequency over recent years,^24^ it has shown a positive association with lactate clearance and survival.^24–28^ However, estimates from meta-analyses evaluating use of PAC in patients with cardiogenic shock are inconsistent,^29–31^ and clinical trials on this intervention are lacking (currently enrolling PACCS trial: [NCT05485376]).^32^ Our finding, although limited by residual confounding, may serve as a signal to revisit this discussion.

Our study has several limitations. First, we used retrospective administrative data with many unmeasured confounders that include lack of vital signs, hemodynamics, medications doses, and other clinical data. Although we used multivariable regressions with available data, there is high risk of residual confounding in our estimates. However, our findings are consistent with those of other datasets, including those of sub-analyses of clinical trials, and persisted in several sensitivity and subgroup analyses. Second, our observations are date-stamped, without time-of-day data available. Although serially ordered, we cannot adjust for time interval in hours between laboratory results. This limitation, however, would move our estimates towards the null, as exemplified in our sensitivity analyses expanding the observation period to 2 days. Third, we used data from mostly academic centers in the US, which may limit generalizability to other scenarios.

In conclusion, in this multicenter observational study, nearly 1 in 3 patients with cardiogenic shock and elevated lactate achieved normal lactate levels on day of admission, which was associated with significant improvement in survival. Across groups of patients with different initial lactate levels, the magnitude of change in lactate concentration during the first day of admission was associated with significant changes in mortality. Furthermore, the association between PAC use and successful lactate clearance warrants further investigation as a potentially modifiable lever to improve outcomes in this high-risk population.

## ACKNOWLEDGEMENTS

We thank Vizient for providing the dataset.

## DATA AVAILABILITY

The data underlying this article were provided by Vizient® Clinical Data Base by permission. Data may be requested directly to Vizient® Clinical Data Base. Code used to analyze the data is publicly available at https://github.com/cesarcaraballoc/cs-lactate-clearance.

## SUPPLEMENTAL MATERIALS

### Supplemental Tables

**Table S1.** Procedure codes.

**Table S2.** Unadjusted and adjusted association between day 1 interventions and early lactate clearance among patients with cardiogenic shock.

### Supplemental Figures

**Figure S1.** Flow chart of study population.

**Figure S2.** Distribution of initial lactate, last lactate, and lactate change among the study population.

**Figure S3.** Heatmap of Observed In-Hospital Mortality Risk Among Patients with Cardiogenic Shock, Relative Change.

**Figure S4.** Association between relative change in serum lactate concentration on day of admission and in-hospital mortality among patients with cardiogenic shock.

**Figure S5.** Heatmap of Observed In-Hospital Mortality Risk Among Patients with Cardiogenic Shock, with Extended Lactate Clearance Period.

**Figure S6.** Association between change in serum lactate concentration within the first two days of admission and in-hospital mortality among patients with cardiogenic shock and initial serum lactate elevation.

**Figure S7.** Heatmap of Observed In-Hospital Mortality Risk Among Patients with Cardiogenic Shock, with Lactate Threshold Increased to 4 mmol/L.

**Figure S8.** Association between change in serum lactate concentration and in-hospital mortality among patients with cardiogenic shock and initial serum lactate elevation greater than 4 mmol/L.

**Figure S9.** Kaplan-Meier survival curves among patients admitted with cardiogenic shock and elevated serum lactate, stratified by lactate clearance status on day of admission

